# Association between maternal breastmilk microbiota composition and rotavirus vaccine response in African, Asian, and European infants: a prospective cohort study

**DOI:** 10.1101/2022.11.09.22282115

**Authors:** Jonathan Mandolo, Edward P. K. Parker, Christina Bronowski, Kulandaipalayam Natarajan C. Sindhu, Alistair C. Darby, Nigel A. Cunliffe, Gagandeep Kang, Miren Iturriza-Gómara, Arox W. Kamng’ona, Khuzwayo C. Jere, the RoVI study group

**Author notes:** Corresponding authors:* Khuzwayo C. Jere. These authors contributed equally (Jonathan Mandolo, Edward P. K. Parker). These authors jointly supervised this work (Gagandeep Kang, Miren Iturriza-Gómara, Arox W. Kamng’ona, Khuzwayo C. Jere).

## Abstract

**Background:** Maternal breastmilk is a source of pre- and pro-biotics that impact neonatal gut microbiota colonisation. Since oral rotavirus vaccines (ORVs) are administered at a time when infants are often breastfed, breastmilk microbiota composition may have a direct or indirect influence on vaccine take and immunogenicity.

**Methods:** Using standardised methods across sites, we compared breastmilk microbiota composition in relation to geographic location and ORV response in cohorts prospectively followed up from birth to 18 weeks of age in India (*n* = 307), Malawi (*n* = 119), and the UK (*n* = 60).

**Results:** Breastmilk microbiota diversity was higher in India and Malawi than the UK across three longitudinal samples spanning weeks of life 1 to 13. Dominant taxa such as *Streptococcus* and *Staphylococcus* were consistent across cohorts; however, significant geographic differences were observed in the prevalence and abundance of common and rare genera throughout follow-up. No significant associations were identified between breastmilk microbiota composition and ORV outcomes including seroconversion, post-dose 1 vaccine shedding, and/or post-vaccination rotavirus-specific IgA level.

**Conclusions:** Our findings suggest that breastmilk microbiota composition may not be a key factor in shaping trends in ORV response within or between countries.

## INTRODUCTION

Maternal breastmilk is a key source of nutrition for newborn infants. It is enriched with a variety of macro- and micro-nutrients vital for infant growth, and contains immunoglobulins, growth hormones, and oligosaccharides that perform critical functions in infant gut homeostasis and immune development [1]. The *Bifidobacteriaceae, Pseudomonadaceae, Streptococcaceae, Enterococcaceae* and *Staphylococcaceae* bacterial families have consistently been identified as core constituents of the breastmilk microbiota [2–4]. These and other bacteria in breastmilk may act as a source of commensal bacteria, seeding the infant gut microbiota at a critical stage of neonatal development [5]. Geographic region, mode of delivery, maternal health, and genetic factors are among the factors associated with maternal breastmilk microbiota composition [2,3,6–9].

Rotavirus remains a major cause of severe gastroenteritis among children worldwide. More than 100 countries have incorporated oral rotavirus vaccine (ORV) into their national immunization programs [10]. Malawi and the UK introduced the live-attenuated monovalent G1P[8] Rotarix vaccine into their national immunisation programs in 2012 and 2013, respectively [11,12]. India introduced a live-attenuated, monovalent vaccine containing a G9P[11] human strain into its immunisation program in 2016 [13]. These vaccines have reduced the burden of rotavirus, although in India and Malawi the estimated mortality burden due to rotavirus remained significant as of 2016 (9.2 and 31.2 per 100,000, respectively, compared to 0.1 per 100,000 in England) [14]. As reported for other live oral vaccines such as oral poliovirus vaccine, ORV immunogenicity and efficacy is significantly reduced in low- and middle-income (LMIC) compared with high-income countries [15]. Given that LMICs account for approximately 95% of all rotavirus deaths worldwide [16], the public health burden associated with impaired ORV response is considerable.

Several mechanisms may contribute to the impaired performance of ORV in LMICs. In Malawi and India, we reported infant gut microbiota diversity to be negatively correlated with ORV response [17,18]. Maternal rotavirus-specific IgG and IgA antibodies in breastmilk and serum were also negatively correlated with ORV response [17,19], although similar correlations were absent among infants in the UK [17]. Other factors which may impact ORV response include histo-blood group antigen status, environmental enteric dysfunction (EED), and pre-vaccination rotavirus exposure [20].

Since ORV is administered at a time when infants are often breastfeeding, we hypothesised that maternal breastmilk microbiota composition may be associated with ORV response, either by directly interacting with the vaccine viruses or indirectly via the developing infant gut microbiota. We tested this hypothesis using standardised methods across cohorts in Malawi, India, and the UK [17,18].

## MATERIALS AND METHODS

### Study cohort

This is a follow-up to the Rotavirus Vaccine Immunogenicity (RoVI) study – a multi-site observational cohort study exploring the impact of maternal antibodies, microbiota development, and EED on ORV response (CTRI/2015/11/006354). The study design, sample handling, lab assays, and primary outcomes of the study have been described previously [17,18]. Briefly, pregnant women were recruited across sites in Blantyre (Malawi), Vellore (India), and Liverpool (UK). Infants received routine immunisations including two doses of Rotarix according to the national immunisation schedule at each study site (weeks of life 6 and 10 in India and Malawi; weeks of life 8 and 12 in the UK). Rotavirus-specific IgA (RV-IgA) was measured in infant blood samples collected pre- and 4 weeks post-vaccination. Rotavirus shedding was measured in six longitudinal stool samples per infant, including 1 week after each ORV dose. Breastmilk samples were collected in week of life 1 and in the week after each ORV dose (**Figure 1A**).

**Figure 1.**
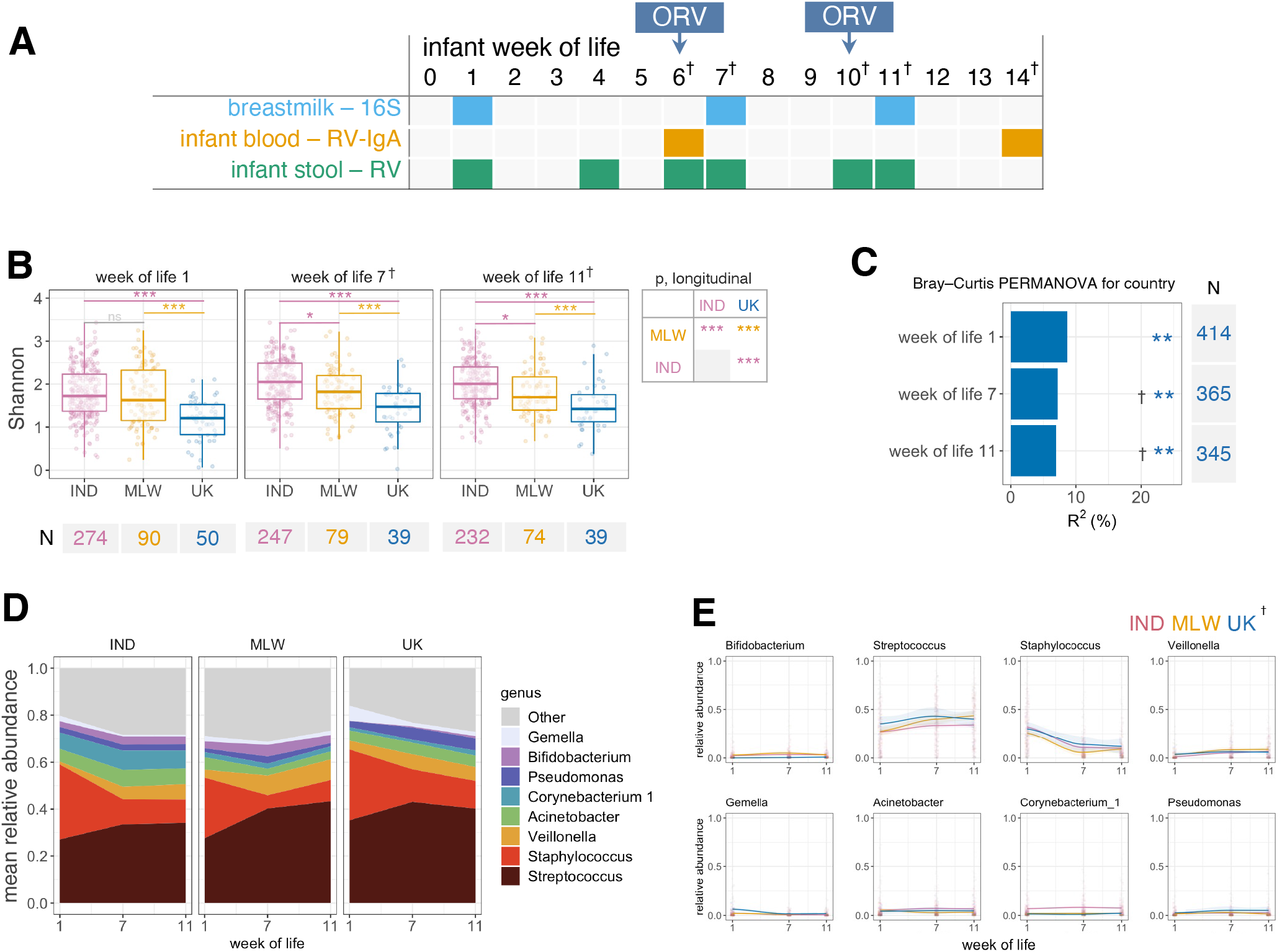
Geographic differences in breastmilk microbiota composition. **(A)** Sample collection strategy. **(B)** Analysis of alpha diversity, based on genus-level Shannon index. Cross-sectional comparisons were performed using ANOVA with post-hoc Tukey tests. Longitudinal comparisons were performed using mixed-effects regressions with false discovery rate correction of pairwise comparisons. **(C)** Proportion of variation in microbiota composition associated with country, calculated via PERMANOVA using genus-level unweighted Bray–Curtis distances. **(D)** Longitudinal plot of mean genus abundances. Genera are included if present with a mean relative abundance of ≥5% in at least one country at one or more timepoints. **(E)** Longitudinal relative abundance plots for major genera by country. Lines show local weighted regression (loess) fits with 95% confidence intervals. IND, India; MLW, Malawi; ns, not significant; †, +2 weeks samples collected at weeks of life 7 and 11 in the UK due to later vaccination schedule; *p < 0.05; **p = 0.001; ***p < 0.0005.

### Sample processing

Breastmilk samples were collected in sterile sample pots by participants and transferred to the site-specific laboratory by courier within 24 hours (and refrigerated throughout) in the UK or within 4 hours in India and Malawi. Upon receipt, samples were kept at 4°C for a maximum of 8 hours until processing and subsequently stored in 2 ml aliquots in SuperLock tubes (Starlab) at −70 °C for a maximum of 2 weeks prior to DNA extraction. DNA was extracted from 1 ml of breastmilk but otherwise followed the stool-specific protocol previously described [17]. A negative extraction control was included in each DNA extraction batch. DNA extracts from Malawi and India were shipped on dry ice to the University of Liverpool for library preparation and sequencing.

### Microbiota sequencing

Breastmilk microbiota composition was determined by sequencing the V3–V4 region of the 16S rRNA gene. Amplicon generation, library preparation, and sequencing steps were performed as previously described for stool [17], but with 15 cycles (as opposed to 10) for the initial amplicon PCR and 20 cycles (as opposed to 15) for subsequent indexing PCR to ensure robust amplification from the low-biomass samples. We sequenced amplicons for 1,301 separate breastmilk samples (894 from India, 275 from Malawi, and 132 from the UK) across 6 Illumina HiSeq2500 lanes (v2 chemistry with 600 cycles in rapid run mode). Samples from each participant were processed on the same plate. Sequencing was batched by geographic location according to sample availability. Each PCR plate included: a no-template PCR control; a breastmilk control sample provided by a mother in the UK who was not enrolled in the study; DNA from a mock community (Zymo Research D6306); and a pool of extraction controls corresponding to the samples contained on each plate for samples from India and the UK. Due to shipment challenges, extraction controls corresponding to 141/243 (58%) of samples from Malawi were included in the extraction pools. To better define the amplicon profile of extraction controls, we sequenced a further 49 pools containing 1–5 controls from extraction batches performed in India or the UK. Final libraries contained up to four 96-well PCR plates (384 amplicons). Breastmilk DNA samples were amplified on separate plates to stool samples, though we allowed mixing of stool and breastmilk PCR plates in a given library. To validate the robustness of the sequencing protocol, 90 breastmilk DNA samples (30 per cohort, all collected in week of life 1) were transferred to Imperial College London and sequenced according to the methods above with minor modifications, as previously described [17].

### Bioinformatic processing

Adapters were trimmed from raw sequences using cutadapt version 1.18 [21]. We merged, filtered, and denoised the amplicon sequences using the DADA2 pipeline in QIIME2 (version 2018.11) [22]. Forward and reverse reads were truncated to 270 bp and 200 bp, respectively. Taxonomic assignment was performed via the *dada2* package (version 1.14.1) using the RDP naïve Bayesian classifier trained on the Silva rRNA database (version 132). Ribosomal sequence variants (RSVs) were retained if they were 390–440 bp in length, assigned as bacterial, detectable at •0.1% abundance in at least one sample, and passed frequency-based contamination filtering using the *decontam* package in R (version 3.6.1) [23]. Nanodrop readings (ng/•l) were used to define concentration of the input template.

Given the additional amplification involved in library preparation for breastmilk samples, reads were frequently detected in extraction controls (*n* = 56 individual or pooled controls with >10,000 reads after the filtering steps above). Several additional filtering steps were therefore included. First, we retained RSVs if they were detectable at •0.1% abundance in •1% of breastmilk samples from at least one country. Second, we applied prevalence-based filtering using the *decontam* package with a *p* value threshold of 0.05 to exclude RSVs that were more common in extraction controls. Finally, we removed samples if their mean Bray-Curtis distance (based on either weighted or unweighted metrics) from breastmilk extraction controls was smaller than their mean distance from other breastmilk samples collected from the same country (**Supplementary Figure 1**).

### Outcomes

We compared breastmilk microbiota composition by country and ORV response. Our primary indicator of ORV response was seroconversion status – defined as a 4-fold increase in RV-IgA concentration or detection of antibodies at •20 IU/ml in previously seronegative infants. Secondary outcomes included post-vaccination RV-IgA concentration (as a continuous variable) and rotavirus shedding 1 week after the first dose of ORV (as an indicator of vaccine virus take). Shedding was detected via real-time PCR targeting the Rotarix *NSP2* gene [24]. We also performed an exploratory analysis of alpha and beta diversity to identify demographic and clinical factors associated with breastmilk composition.

### Statistical analysis

Analyses were performed in the programming language R following the statistical pipeline previously described for stool samples with minor modifications [17]. Alpha and beta diversity were calculated at a rarefaction depth of 15,000 sequences per sample. We performed cross-sectional analyses of alpha diversity via analysis of variance (ANOVA), logistic regression (binary ORV outcomes), Pearson’s r with two-sided hypothesis testing (log-transformed RV-IgA), and linear regression (exploratory covariates). We assesses beta diversity using permutational multivariate ANOVA (PERMANOVA) with 999 permutations based on genus-level unweighted Bray–Curtis distances. For binary outcomes, discriminant genera and RSVs were identified via two-sided Fisher’s exact test (differences in prevalence) and Aldex2 (two-sided Wilcoxon rank-sum test of centred log-ratio transformed sequence counts), with taxa classified as discriminant if they had a *p* value of <0.05 based on either method after Benjamini–Hochberg false discovery rate (FDR) adjustment. Aldex2 was used to identify taxa correlated with log-transformed RV-IgA (FDR-adjusted *p* value of <0.05 based on two-sided Spearman’s rank test). Taxa were included if they were detected with a prevalence of >5% in at least one of the groups being compared. We supplemented cross-sectional analyses with longitudinal mixed-effects models of Shannon index and taxon abundances (zero-inflated negative binomial models of genus-level sequence counts), including week of life as a covariate and study ID as a random effect. Genera were included in longitudinal models if they were present in 20% of samples in a given country.

We applied Random Forests in a series of cross-sectional analyses to predict country and ORV outcome based on genus or RSV relative abundances. For each analysis, we performed 20 iterations of 5-fold cross-validation. For binary outcomes, we standardised the baseline accuracy of classification models at 50% by fitting each iteration of cross-validation on a random subset of 50 samples per group (or the number of samples in the minority group if this was <50). Models were excluded if there were <10 samples in the minority group. For regression models, accuracy was quantified by using linear regression to determine the out-of-bag R^2^ values for predicted vs observed RV-IgA values.

For positive controls and technical replicates, we used linear regression (alpha diversity and common genera) and PERMANOVA (beta diversity) to quantify the proportion of variance explained by sample ID.

The raw sequence data for this study have been deposited in the European Nucleotide Archive under accession code PRJEB38948. Processed data and analysis code are available on Github (https://github.com/eparker12/RoVI).

### Ethics approval

The study was approved by the Institutional Review Board at the Christian Medical College (CMC) in Vellore (IRB No. 9472/24.06.2015), the College of Medicine Research and Ethics Committee in Blantyre (P.01/16/1853), and the North West—Liverpool Central Research Ethics Committee in Liverpool (15/NW/0924).

## RESULTS

### Study cohort

Overall, 664 mother–infant pairs (395 in India, 187 in Malawi, and 82 in the UK) were enrolled in the study and the primary endpoint (measurement of seroconversion or dose 1 shedding) was reached for 484 (307 in India, 119 in Malawi, and 60 in the UK). Baseline characteristics, wild-type rotavirus infection status, EED biomarker levels, and infant stool microbiota composition have previously been compared by country and ORV outcome [17]. Exclusive breastfeeding was reported by 265/307 (86%) mothers in India, 108/119 (91%) in Malawi, and 26/60 (43%) in the UK, with partial breastfeeding reported by a further 32/307 (10%) in India, 11/119 (9%) in Malawi, and 20/60 (33%) in the UK. Exclusive breastfeeding was positively correlated with ORV seroconversion and post-vaccination infant RV-IgA levels in India but not in other cohorts. Breastmilk RV-IgA levels were negatively correlated with infant RV-IgA levels in India and Malawi [17].

### ORV shedding and immunogenicity

As previously reported [17], seroconversion was observed in 27/51 (53%) infants in the UK, 85/305 (28%) in India, and 24/103 (23%) in Malawi. Rotavirus shedding 1 week after the first dose of ORV was detected in 55/60 (92%) infants in the UK, 82/305 (27%) in India, and 56/101 (55%) in Malawi. Geometric mean concentrations (GMCs) of RV-IgA (IU/ml) after vaccination were 27 (17–45) in the UK, 20 (95% CI 16–25) in India, and 9 (6–12) in Malawi.

Indian infants were characterised by high rates of neonatal rotavirus infection, defined as detection of wild-type rotavirus shedding in week 1 of life or baseline seropositivity (pre-vaccination RV-IgA •20 IU/ml). This was observed in 166/304 (55%) infants in India, 10/90 (11%) in Malawi, and 2/54 (4%) in the UK. Given the potential impact of neonatal infection on ORV shedding and immunogenicity [17], we report results for the Indian cohort overall and stratified by neonatal infection status below.

### Geographic differences in breastmilk microbiota composition

Of 1,301 breastmilk samples sequenced from this study population, 1,124 yielded high-quality microbiota profiles (•15,000 sequences after quality filtering; 95,075 ± 113,894 [mean±s.d.] sequences per sample). Microbiota profiles of positive controls and technical replicates were consistent across sequencing runs and facilities (**Supplementary Figure 2**).

There were marked differences in breastmilk microbiota composition between cohorts. Microbiota diversity was significantly lower in UK than both other cohorts at all timepoints. Diversity was similar in India and Malawi at week of life 1, but higher in India than Malawi at weeks of life 7 and 11 (**Figure 1B**). Samples clustered by individual (PERMANOVA R^2^ = 49%, p = 0.001), with country accounting for 6–9% of variation depending on age (**Figure 1C**). Although 350 genera were detected overall, a small proportion were dominant in each cohort (**Figure 1C** and **Supplementary Figure 3**). Among dominant genera, *Streptococcus* was depleted in India compared with both other cohorts, while *Acinetobacter* and *Corynebacterium* were enriched. *Staphylococcus* followed a parallel trajectory in each cohort, peaking in week of life 1, and was less abundant in Malawi than both other cohorts. *Bifidobacterium* was observed at lower abundance in the UK, reflecting the pattern previously reported for stool samples [17], while *Gemella* was enriched in this cohort (**Figure 1D**).

Additional discriminant taxa were identified when considering both common and rare genera via longitudinal and cross-sectional models (**Supplementary Figure 4**). Based on longitudinal models, 17 genera were enriched in India compared with both other cohorts, including nine Proteobacteria (e.g. *Aeromonas* and *Alishewenalla*), three Firmicutes (e.g. *Enterococcus* and *Aerococcus*) and five Actinobacteria (e.g. *Dermacoccus*). Nine genera were enriched in Malawi compared with both other cohorts, including the Bacteroidetes genus *Prevotalla* 9 alongside eight Firmicutes (e.g. *Faecalibacterium* and *Lachnospiraceae*). Three genera – *Gemella, Haemophilus*, and *Enterobacter* – were enriched in the UK compared with the other cohorts.

Random Forests discriminated samples by country with high accuracy based on genus relative abundance (median cross-validation accuracies of 85–95%; baseline accuracy 50%; **Supplementary Figure 5**). Genera underlying the predictive accuracy (based on mean importance scores) were consistent with the discriminant taxa described above (**Supplementary Table 1**).

We also assessed alpha and beta diversity of breastmilk samples in relation to individual-level variables measured in each cohort (**Figure 2**). With the exception of infant serum •1 acid glycoprotein level (a marker of systemic inflammation), which was modestly associated with beta diversity in Malawian samples (R^2^ 3.7%), no covariates were significantly associated with breastmilk microbiota composition.

**Figure 2.**
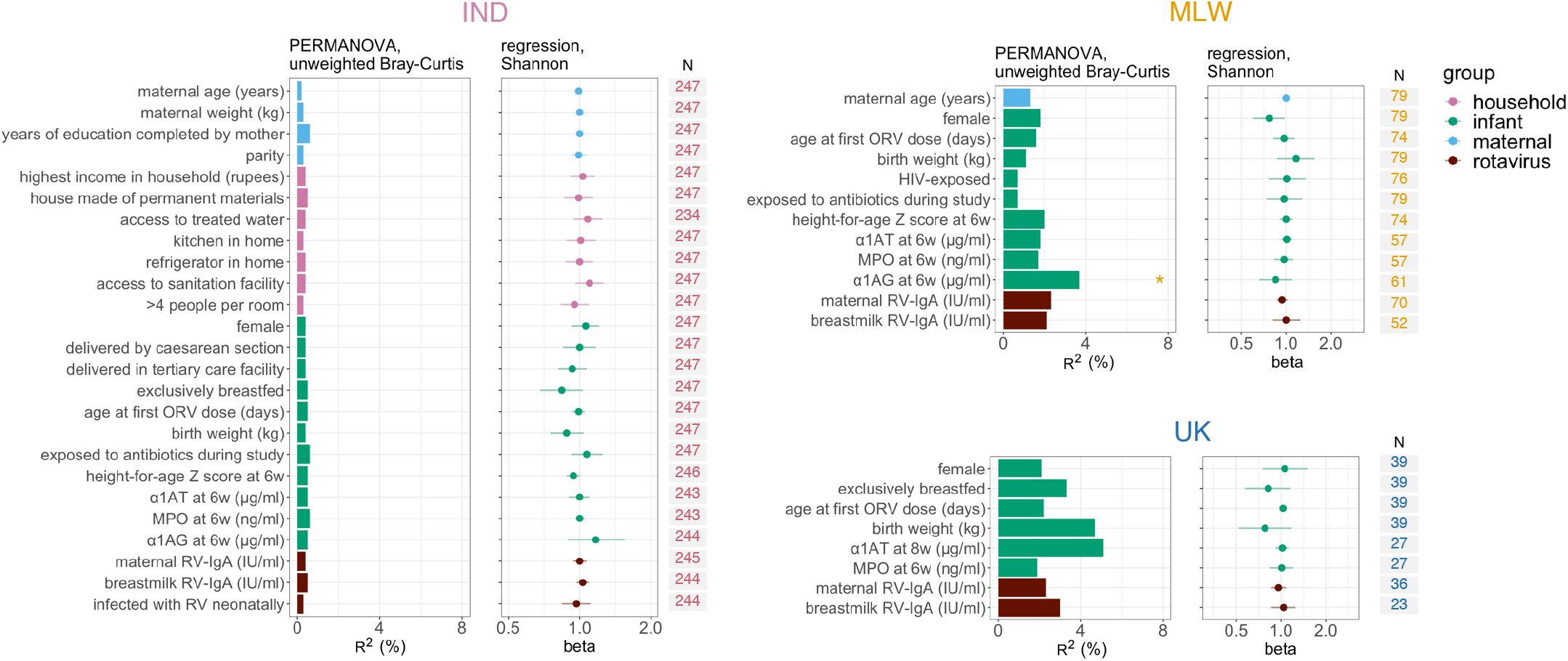
Cofactors associated with breastmilk microbiota composition. Samples collected 1 week after the first dose of oral rotavirus vaccine were included (week of life 7 in India and Malawi; week of life 9 in the UK). The left panel, presenting data for Indian samples (n = 247), contains the full list of exploratory variables (with the exception of HIV exposure status, which was also assessed for Malawi). For analyses of samples from Malawi and the UK (right panels; n = 79 and 39, respectively), variables were excluded if they were not measured or exhibited limited variability (n<10 in either comparison group). PERMANOVA was performed using genus-level unweighted Bray–Curtis distances. Shannon index was calculated at genus level and assessed as an outcome variable via linear regression. α1AT, α1-antitrypsin; α1AG, α1 acid glycoprotein; IND, India; MLW, Malawi; MPO, myeloperoxidase; ORV, oral rotavirus vaccine; RV, rotavirus; * FDR p <0.05.

### Breastmilk microbiota composition versus ORV response

Based on longitudinal models of Shannon index, we observed no significant differences in microbiota diversity according to seroconversion status in any cohort (**Figure 3A**). This was also the case for cross-sectional analyses, with the exception of comparisons in Malawi at week 7 of life (the week after the first dose of ORV), wherein Shannon index was negatively correlated with seroconversion. Beta diversity analyses based on genus-level unweighted Bray-Curtis distances did not reveal any significant association between breastmilk microbiota composition and seroconversion status (**Figure 3B**). Likewise, Random Forest models based on genus or RSV abundances failed to accurately predict seroconversion (**Figure 3C**), and no discriminant taxa were identified based on cross-sectional analyses of prevalence or abundance after FDR correction. Longitudinal models of common genera (•20%) revealed frequent age-associated changes in taxon abundance but only one significant association with seroconversion (a negative correlation between *Alloprevotella* abundance and seroconversion in Malawi; **Supplementary Table 2**).

**Figure 3.**
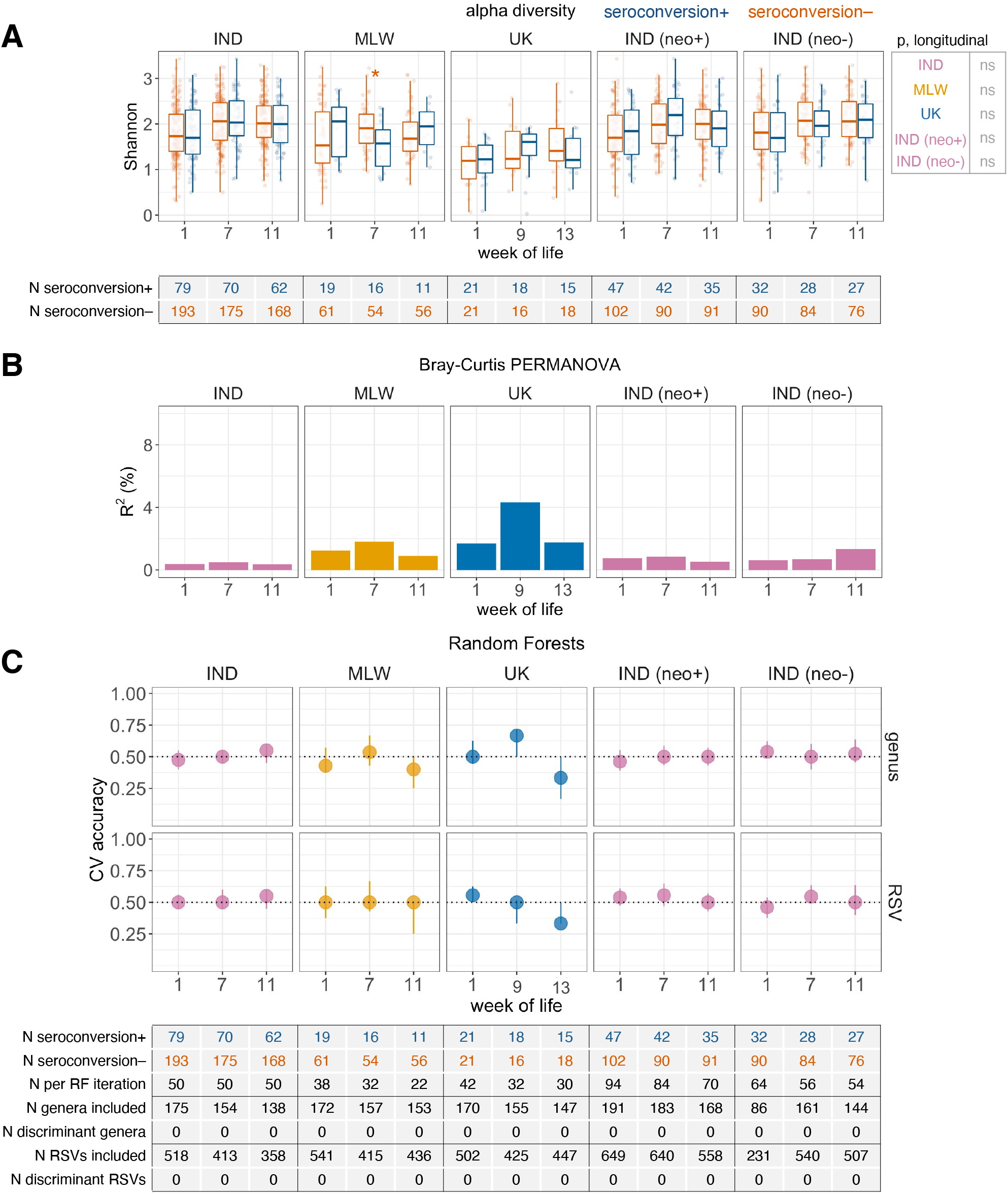
Association between breastmilk microbiota composition and oral rotavirus vaccine seroconversion. **(A)** Analysis of alpha diversity, based on genus-level Shannon index. Cross-sectional comparisons were performed using logistic regression. Longitudinal comparisons were performed using mixed-effects models. (B) Proportion of variation in microbiota composition associated with seroconversion, calculated via PERMANOVA using genus-level unweighted Bray–Curtis distances. (C) Cross-validation accuracy of Random Forests for prediction of seroconversion. Median out-of-bag accuracy (proportion correctly assigned) and interquartile range across 20 iterations of 5-fold cross-validation are displayed. Each iteration included an equal number of responders and non-responders (50 per group where possible, or else the number in the minority group if this was <50). Taxa were classified as discriminant if they had an FDR-adjusted p value of <0.05 based on either two-sided Fisher’s exact test (differences in prevalence) or Aldex2 with two-sided Wilcoxon rank-sum test (differences in abundance). CV, cross-validation; IND, India; MLW, Malawi; neo+, infected with rotavirus neonatally (defined by detection of rotavirus shedding in week of life 1 or baseline seropositivity); neo−, uninfected with rotavirus neonatally; ns, not significant; RF, Random Forests; RSV, ribosomal sequence variant; *p < 0.05.

Cross-sectional analyses of secondary ORV endpoints, including post-vaccination RV-IgA concentration (**Supplementary Figure 6**) and dose 1 ORV shedding (**Supplementary Figure 7**) were consistent with those for seroconversion, revealing no significant associations. Very few discriminant genera were identified with respect to secondary outcomes based on longitudinal models of genus abundance (**Supplementary Table 2**).

## DISCUSSION

Breastmilk is a key source of pre- and pro-biotics that shape infant gut microbiota configuration. This, in turn, plays a pivotal role in shaping immune development. We documented significant differences in breastmilk microbiota composition between Malawi, India, and the UK across the first 13 weeks of life. However, no consistent differences in breastmilk microbiota composition were observed with respect to ORV response.

Despite the geographic differences in breastmilk microbiota composition, several genera of bacteria were dominant across the three cohorts. Together, *Streptococcus, Staphylococcus, Acinetobacter, Bifidobacterium, Veillonella, Gemella, Corynebacterium* and *Pseudomonas* formed approximately 75% for the breastmilk microbiota as determined by 16S amplicon sequencing – consistent with the dominant taxa reported in previous studies [2–4,25]. The relative abundances of these dominant genera changed over time, with *Staphylococcus* declining in abundance from week 1 of life onwards, while *Streptococcus* and *Veillonella* increased in abundance. This is similar to the trajectory in breastmilk microbiota composition reported in Kenya [26]. The infant salivary microbiota is known to be colonised by *Streptococcus* [27,28], such that the continued dominance of *Streptococcus* in maternal breastmilk may partly reflect breastmilk–saliva interplay throughout early life. Skin-associated genera including *Staphylococcus* and *Corynebacterium* were also among the dominant genera in maternal breastmilk, consistent with previous findings [29].

Breastmilk microbiota diversity was higher in Malawi and India compared with the UK. This contrasts with discrepancies we reported in stool microbiota diversity, which was significantly higher in Malawi than both India and the UK at week 1 of life but converged over the ensuing 6–8 weeks [17]. In a previous cross-sectional study spanning 11 study sites, breastmilk microbiota diversity was highest in rural Ethiopia and lowest in Ghana, with intermediate levels across other sites in Africa, Europe, North America, and South America [8]. At genus level, *Streptococcus* was more abundant in Malawi and the UK than in India, while *Bifidobacterium* was depleted in the UK compared with both other cohorts. Prior studies have also highlighted geographically distinct abundance profiles including depletion of *Bifidobacterium* in European compared with African samples [8]. Together, these studies highlight the significant regional variation that occurs in breastmilk microbiota diversity and composition. To delineate overarching global trends (e.g. urban vs rural, high-income vs LMIC), future studies integrating well-powered representative data from multiple countries, such as the present, are warranted.

We did not observe consistent associations between breastmilk microbiota composition and ORV response. At the time of the first dose of ORV, breastmilk microbiota diversity in Malawi was negatively correlated with ORV seroconversion – a correlation that was also apparent among infant stool samples in this cohort [17]. However, while we reported consistent correlations between diversity and seroconversion among Indian and Malawian infants’ stool samples, there was no consistent discrepancy across cohorts in breastmilk. A previous study in India documented higher *Enterobacter/Klebsiella* abundance in breastmilk and infant stool samples of infants with symptomatic rotavirus disease compared to those with asymptomatic or no infection [30]. However, no significant discrepancies were observed between neonates with asymptomatic infection and those lacking infection, which is consistent with the lack of association reported here in relation to attenuated viral exposure via ORV.

To our knowledge, this is the first study to explore the link between breastmilk microbiota composition and ORV response. Our study is strengthened by the use of standardised methods across cohorts, and the exploration of multiple indicators of ORV response, including dose 1 shedding. Nonetheless, several limitations of the present study should be considered. Owing to recruitment challenges in Malawi [17], we fell short of the target sample size in this cohort (*n* = 119 rather than 150), potentially undermining our ability to detect important associations between breastmilk microbiota composition and ORV response in this cohort. Because of their low biomass, breastmilk samples were subjected to extra rounds of PCR amplification to attain adequate material for sequencing (35 cycles vs 25 used for stool), leading to amplification from extraction controls. We accounted for this via stringent abundance- and prevalence-based filtering of potential contaminants and excluded samples which clustered among extraction controls rather than other breastmilk samples. Nonetheless, the potential contribution of contamination and site-specific batch effects to the observed trends cannot be discounted.

Our findings suggest that breastmilk microbiota composition may not be a key factor shaping trends in ORV response within or between countries. Other components of human milk were not considered here and would be a valuable focus of future investigation. Human milk oligosaccharides such as lacto-*N*-tetraose have previously been linked with symptomatic rotavirus infection in Indian neonates, possibly via an effect on neonatal G10P[11] rotavirus infectivity [30]. Future studies of the breastmilk metabolome may help discern whether similar factors influence the immunogenicity and efficacy of ORV.

## Data Availability

The datasets generated during and/or analyzed during the current study are available from the corresponding author following reasonable request.

## Supplementary Materials

Supplementary Figures S1 – S3 and Supplementary Table S1 – S7 are attached.

## Notes

### Disclaimer

The funders had no role in the study design, data collection and interpretation, or the decision to submit the work for publication. The authors received no financial support or other form of compensation related to the development of the manuscript. N.A.C. and K.C.J are affiliated with the National Institute for Health and Care Research (NIHR) Health Protection Research Unit in Gastrointestinal Infections at the University of Liverpool, a partnership with the UK Health Security Agency (UKHSA), in collaboration with the University of Warwick. The views expressed are those of the author(s) and not necessarily those of the NIHR, the Department of Health and Social Care or the UKHSA.

## Acknowledgements

We thank all members of the clinical study teams in Vellore, Blantyre, and Liverpool, including Falak Diab, Siobhan Holt, and the research midwives at the Liverpool Women’s Hospital; Dawn Redman and the team of research nurses at Alder Hey Children’s Hospital; Uma Raman, Charlet, Margaret, Jacklin, and the field research assistants at Christian Medical College, Vellore; and James Tamani, Anna Ainani, Amisa Chisale, Bertha Masamba, Carlo Gondwe, and Evelyn Gondwe in Blantyre, Malawi. Richard Eccles, Anita Lucaci, Richard Gregory, John Kenny, and other staff at the Centre for Genomic Research (University of Liverpool) provided valuable support for the 16S microbiota sequencing work. Above all, we are grateful to the families involved in the study.

## Author contributions

Conceptualisation, M.I.G., A.C.D., E.P.K.P., K.C.J., A.W.K., and G.K.; Methodology, J.M., C.B., E.P.K.P., A.C.D., M.I.G., and G.K.; Software, E.P.K.P.; Validation, E.P.K.P.; Formal Analysis, J.M. and E.P.K.P.; Investigation, J.M., C.B., E.P.K.P., A.W.K., and K.C.J.; Data Curation, C.B., E.P.K.P., and J.M.; Writing – Original Draft, J.M. and E.P.K.P.; Writing – Review & Editing, A.C.D., N.A.C., M.I.G, A.W.K. and K.C.J.; Visualisation, E.P.K.P and J.M.; Supervision, N.A.K. and K.J.; Project Administration, C.B., M.I.G., K.N.S., K.C.J., and G.K.; Funding Acquisition, M.I.G., and G.K. All authors read and approved the manuscript.

## Conference presentation

Part of this work was presented at the 14^th^ International dsRNA Virus Symposium 2022, Banff, Alberta, Canada, 10^th^ October - 14^th^ October 2022 and the 1^st^ Kamuzu University of Health (KUHeS) Research Dissemination Conference, Blantyre, Malawi, 24th - 25th November 2022

---

**Supplementary Figure 1.**
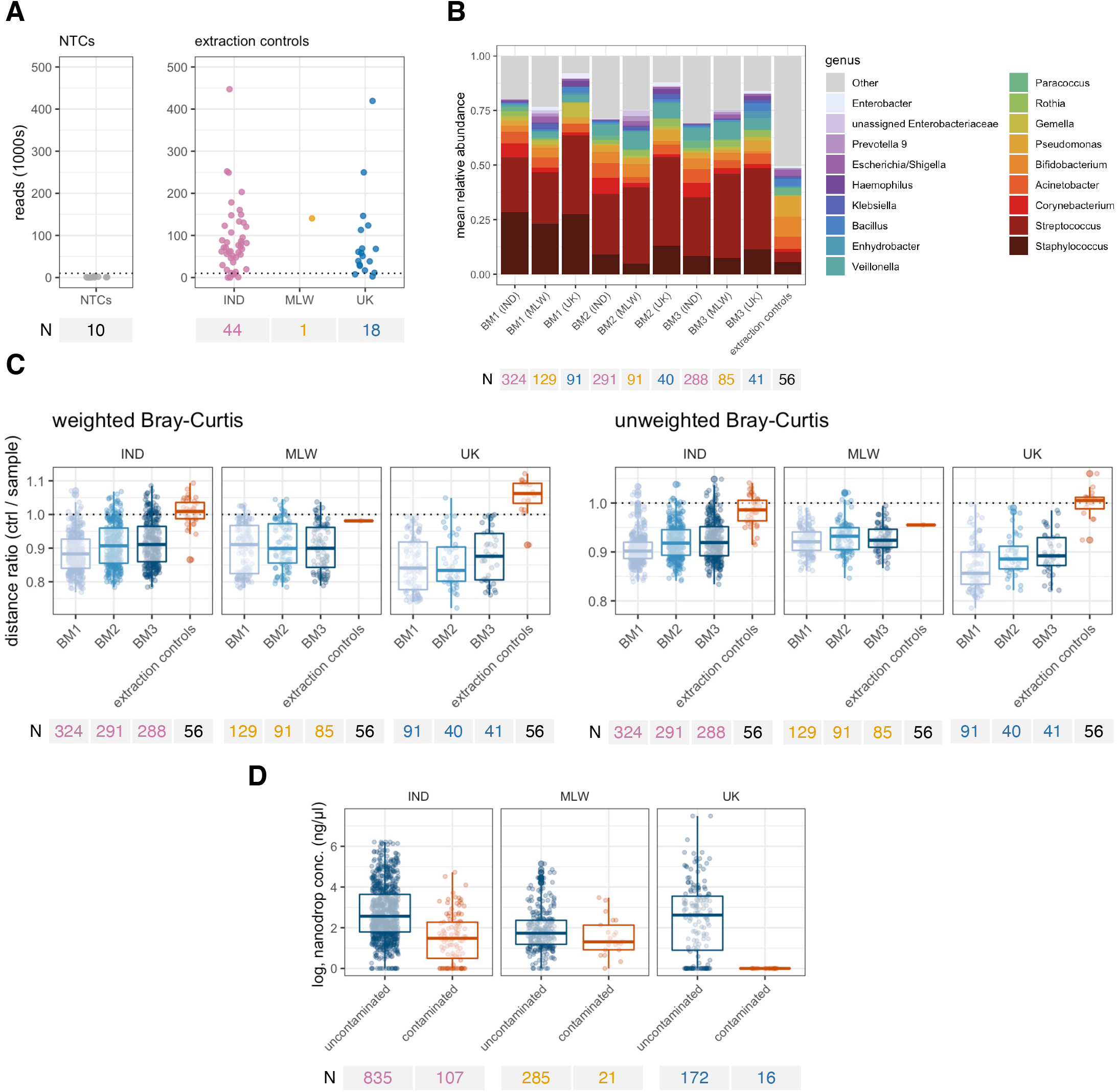
Contaminant filtering steps for breastmilk samples. **(A)** Read counts for negative controls. Counts from were consistently above 10,000 (dotted line) for pooled or individual extraction controls but not NTCs. **(B)** Mean genus abundance profile by sample type and country. Extraction controls displayed a distinct genus abundance profile with notable enrichment of rare taxa (labelled ‘other’). Samples or controls with at least 10,000 sequences were included. **(C)** Identification of samples with a contaminant profile. For each sample, the mean distance was calculated from other breastmilk samples from the same country and all breastmilk extraction controls. If the sample clusters more closely with other samples on average, the ratio of these distances will be <1. **(D)** Nanodrop concentrations of samples identified as potentially contaminated based on either weighted or unweighted Bray-Curtis (ratio >1 in panel [**C**]). A pseudocount of 1 was added before log transformation.

**Supplementary Figure 2.**
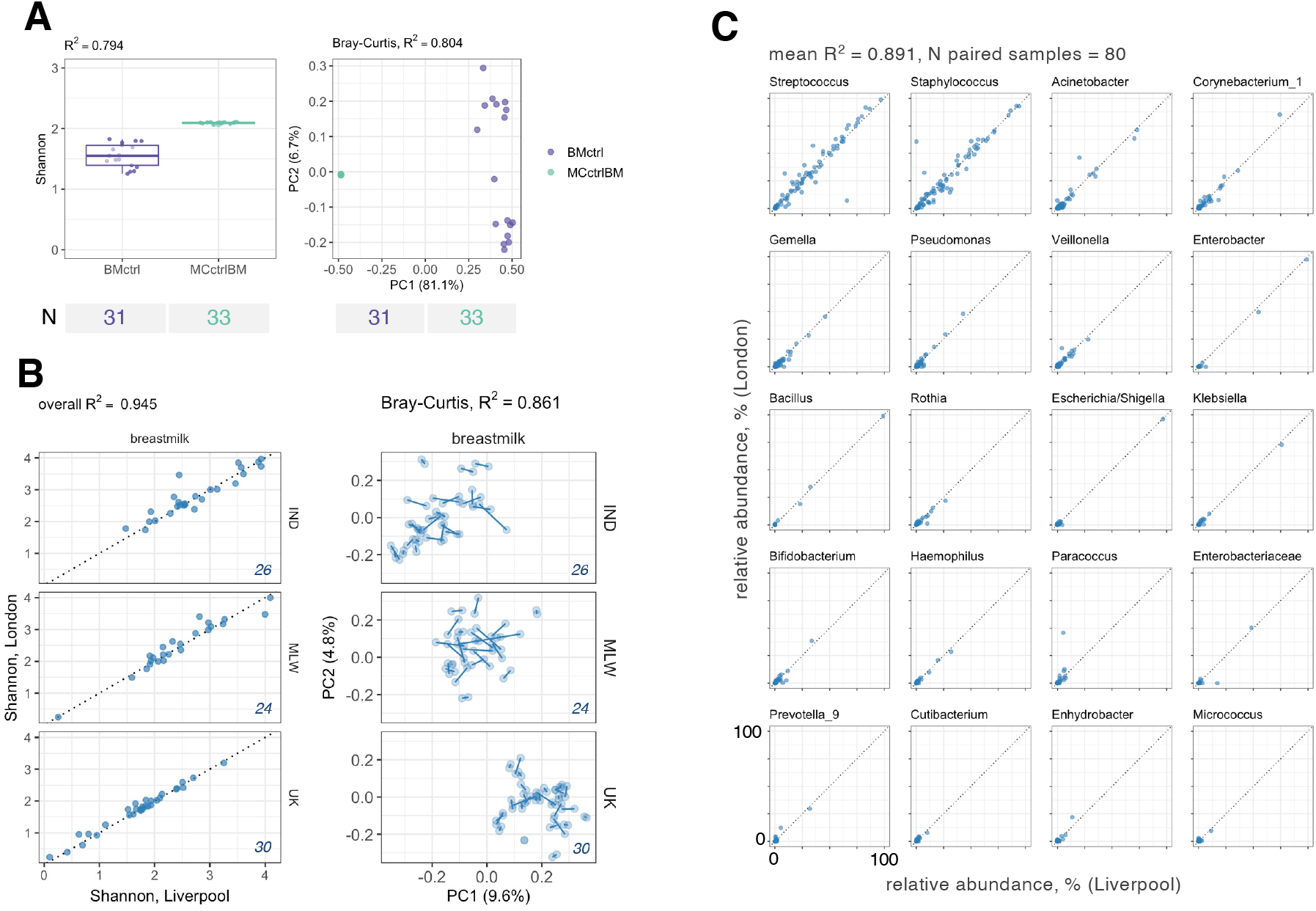
Technical replicate profile. Alpha diversity and beta diversity for **(A)** positive controls and **(B)** validation replicates at ribosomal sequence variant level. Positive controls were included on each PCR plate and included a breastmilk sample (BMctrl) and a mock bacterial community (MCctrl). Validation samples were processed at a separate sequencing facility. These were evenly distributed across the study sites (30 per site per sample group) and randomised across a single sequencing plate. Sample pairs were retained in the analysis of both technical replicates had ≥15,000 sequences after quality filtering (80/90 [89%]). The proportion of variation attributable to sample type was determined by linear regression (for Shannon index) and PERMANOVA of unweighted Bray–Curtis distances (for beta diversity). In **(B)**, technical replicates are linked by a line in the right-hand panel. Sample counts are indicated in italics. **(C)** Relative abundances of major genera in validation samples. R^2^ was determined based on linear regression.

**Supplementary Figure 3.**
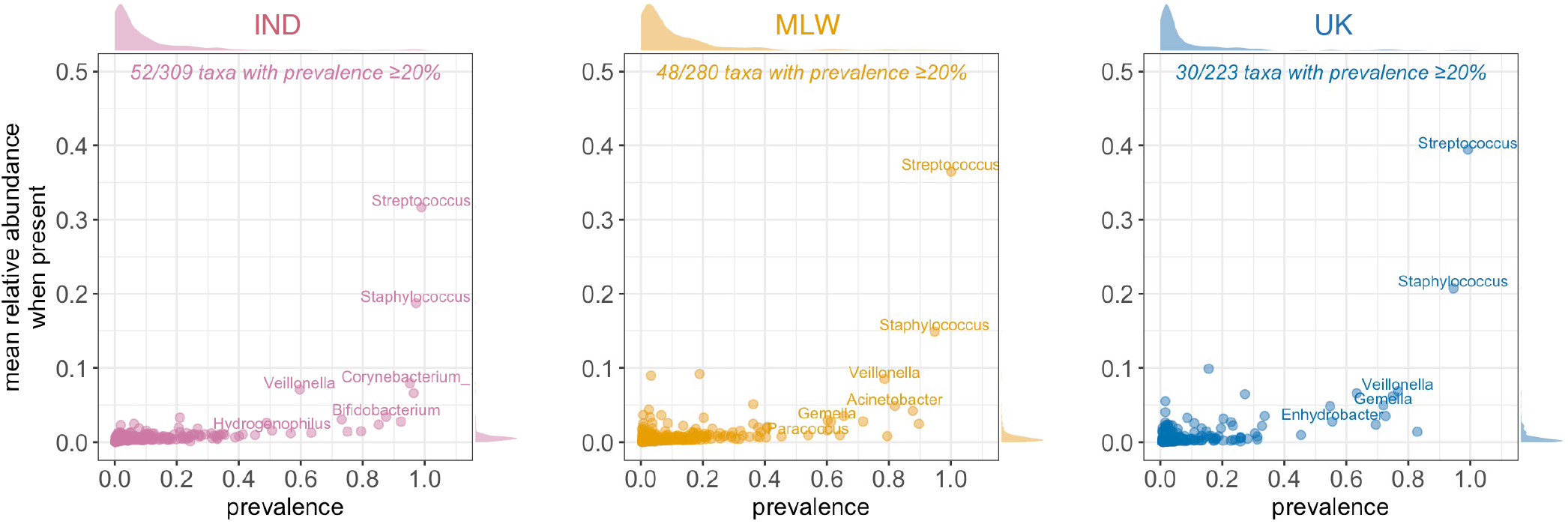
Genus abundance and prevalence profile of breastmilk samples. All breastmilk samples from each cohort were included in the prevalence and abundance calculations (n = 753, 243, and 128 for India, Malawi, and the UK, respectively). Margins display density plots. IND, India; MLW, Malawi.

**Supplementary Figure 4.**
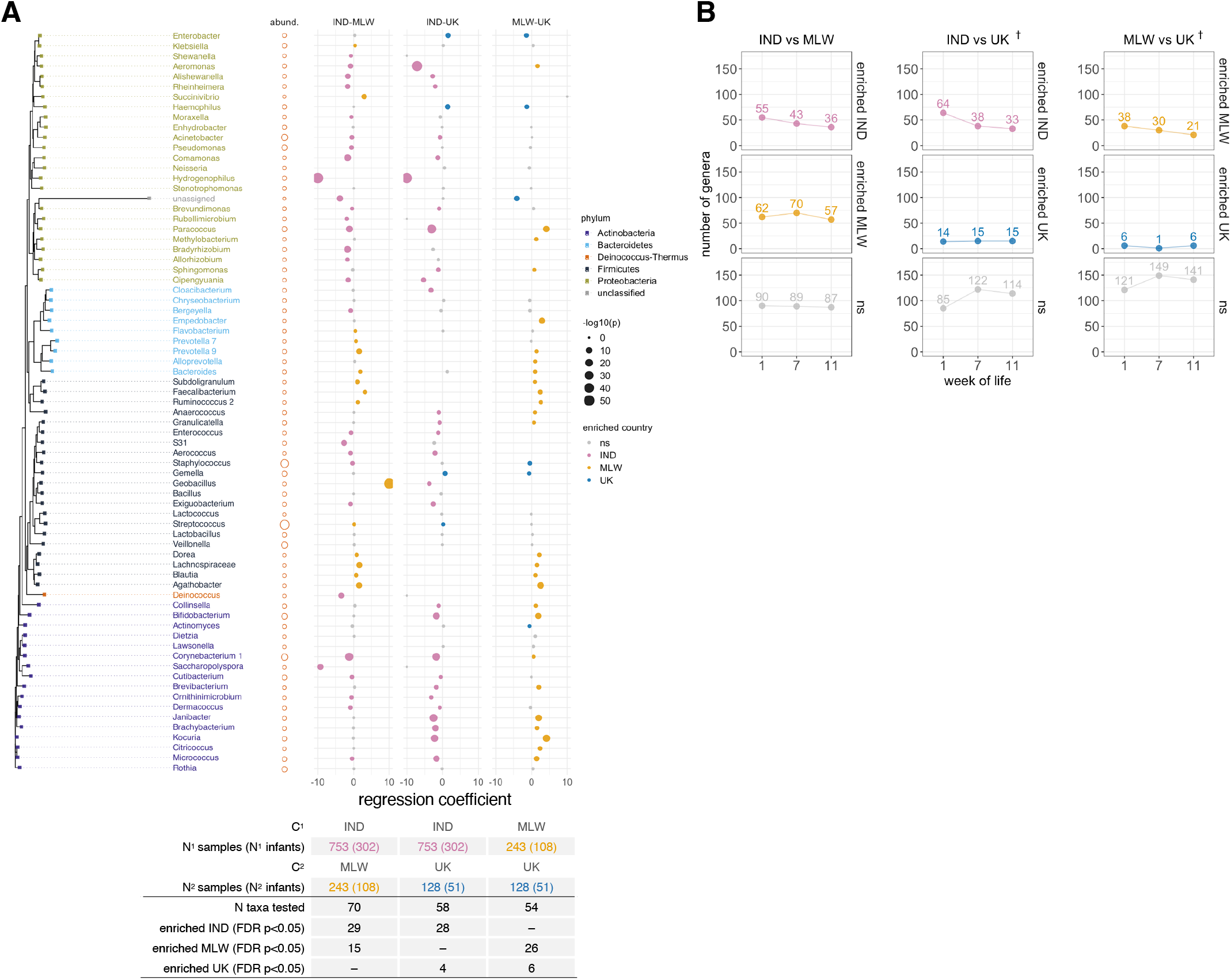
**(A)** Longitudinal models of genus abundance by country. Mixed-effects zero-inflated negative binomial models regressions were used to identify discriminant genera. Genera were included if present in at least 20% of samples from at least one country being compared. Regression coefficients are displayed with point size scaled by p value. Genus order is based on a neighbour-joining tree derived from JC69 distances, with the most abundant ribosomal sequence variant serving as the reference sequence for each genus. Circles to the right of the tree are scaled by mean relative abundance across infant samples (following arcsine square root transformation). **(B)** Cross-sectional comparisons of genus abundance by country. Discriminant genera were identified based on two-sided Fisher’s exact test (differences in prevalence) and Aldex2 with two-sided Wilcoxon rank sum test (differences in abundance). The number of genera with an FDR-adjusted p value of <0.05 based on either method is highlighted for each pairwise cross-sectional comparison. C, country; FDR, false discovery rate; IND, India; MLW, Malawi; ns, not significant; †, +2 weeks samples collected at weeks of life 7 and 11 in the UK due to later vaccination schedule. See **Supplementary Table 1** for full details of discriminant taxa.

**Supplementary Figure 5.**
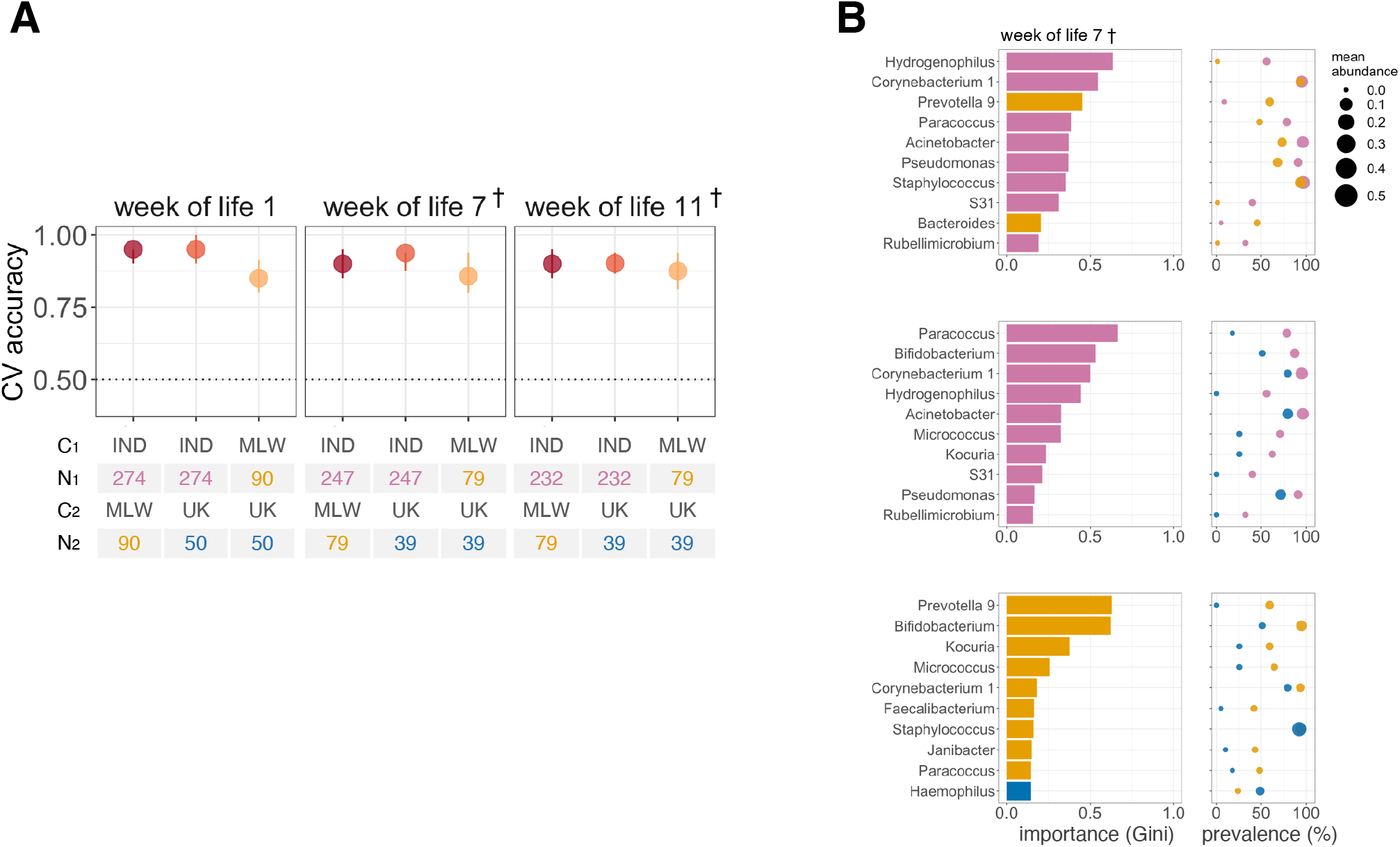
Prediction of country by Random Forests. **(A)** Cross-validation accuracy of Random Forests. Median out-of-bag accuracy (proportion correctly assigned) and interquartile range across 20 iterations of 5-fold cross-validation are displayed. A random subset of 50 samples per country was used for each iteration. (**B**) The 10 most important genera selected by Random Forests for discriminating infants by country in the week after the first dose of oral rotavirus vaccine. Mean cross-validation importance scores based on Gini index are depicted alongside the prevalence and mean abundance of the corresponding genera. C, country; CV, cross-validation; IND, India; MLW, Malawi; †, +2 weeks in UK due to later vaccination schedule.

**Supplementary Figure 6.**
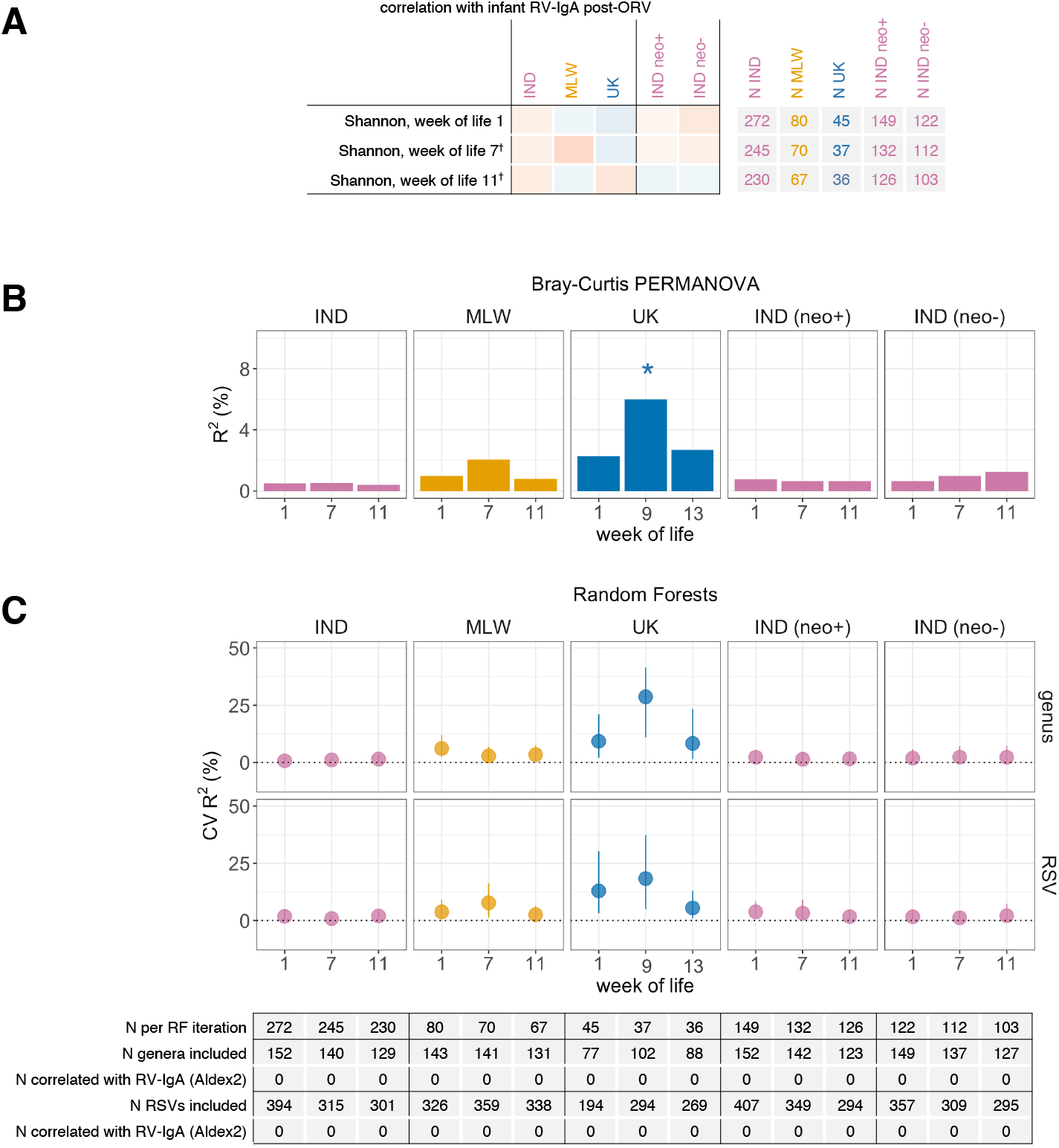
Association between breastmilk microbiota composition and post-vaccination rotavirus-specific IgA concentration. **(A)** Analysis of alpha diversity, based on genus-level Shannon index. Shannon index was compared with log-transformed RV-IgA values using Pearson’s correlation coefficient (*r*) with two-sided hypothesis testing. **(B)** Proportion of variation in microbiota composition associated with RV-IgA, calculated via PERMANOVA using genus-level unweighted Bray–Curtis distances. **(C)** Cross-validation accuracy of Random Forests for prediction of post-vaccination RV-IgA. Median out-of-bag R^2^ and interquartile range are displayed for predicted vs observed RV-IgA across 20 iterations of 5-fold cross-validation. Correlations between log-ratio transformed taxon abundance counts and RV-IgA were determined via Aldex2 with two-sided Spearman’s rank test. Taxa were classified as discriminant if they had an FDR-adjusted p value of <0.05. IND, India; MLW, Malawi; neo+, infected with rotavirus neonatally (defined by detection of rotavirus shedding in week of life 1 or baseline seropositivity); neo−, uninfected with rotavirus neonatally; ns, not significant; RF, Random Forests; RSV, ribosomal sequence variant; * p < 0.05.

**Supplementary Figure 7.**
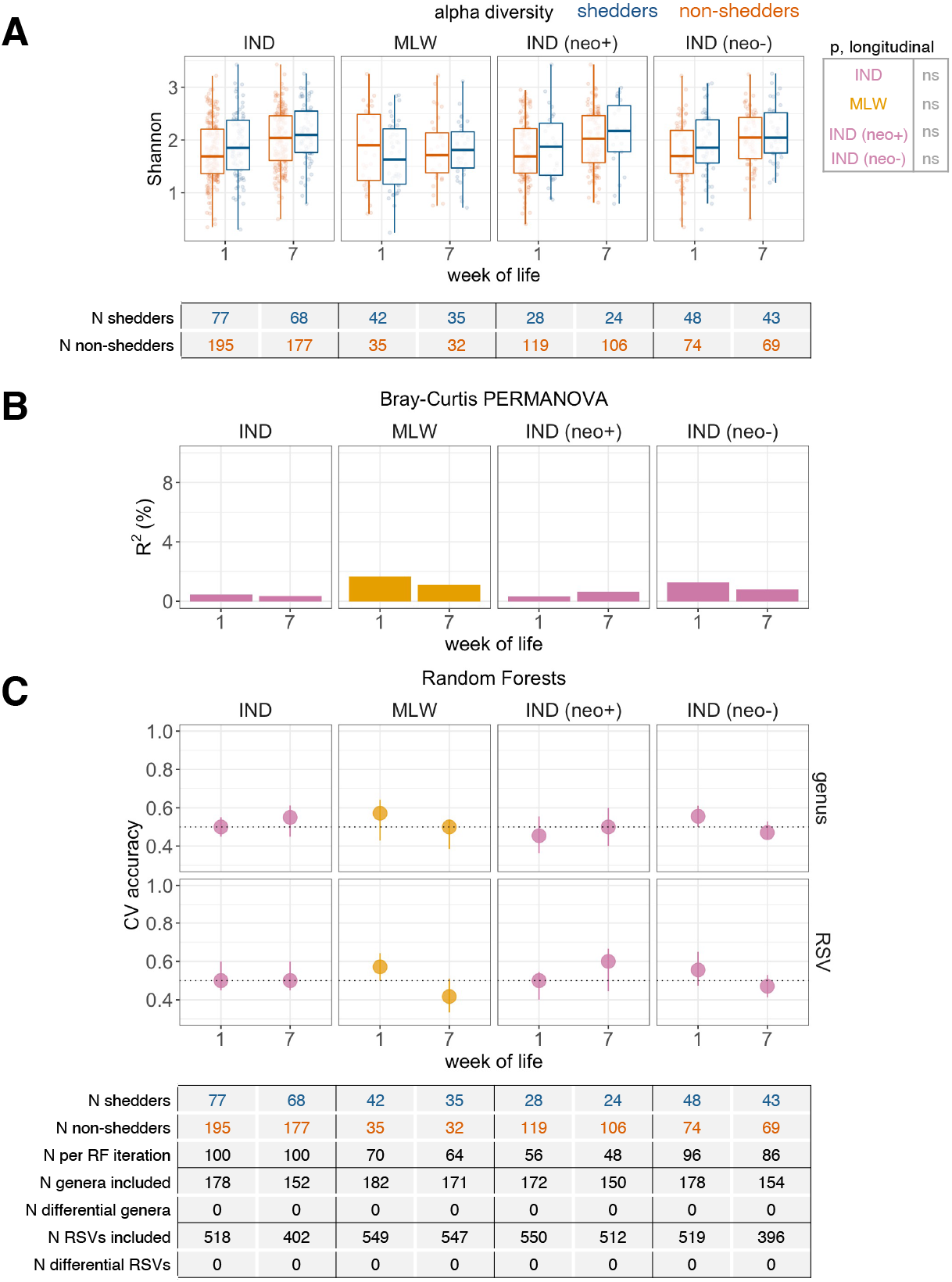
Association between breastmilk microbiota composition and dose 1 oral rotavirus vaccine shedding. See **Figure 3** for details; the same analyses of (A) alpha diversity, (B) beta diversity, and (C) Random Forests cross-validation accuracy are presented here with shedding 1 week after the first dose of oral rotavirus vaccine as outcome. Comparisons were not performed for the UK due to the small number of non-shedders (5 out of 60 infants) in this cohort. CV, cross-validation; IND, India; MLW, Malawi; neo+, infected with rotavirus neonatally (defined by detection of rotavirus shedding in week of life 1 or baseline seropositivity); neo−, uninfected with rotavirus neonatally; ns, not significant; RF, Random Forests; RSV, ribosomal sequence variant; *p < 0.05.

